# Genetic and Phenotypic Evidence for the Causal Relationship Between Aging and COVID-19

**DOI:** 10.1101/2020.08.06.20169854

**Authors:** Kejun Ying, Ranran Zhai, Timothy V. Pyrkov, Marco Mariotti, Peter O. Fedichev, Xia Shen, Vadim N. Gladyshev

## Abstract

Epidemiological studies revealed that the elderly and those with co-morbidities are most susceptible to COVID-19. To understand how genetics affects the risk of COVID-19, we conducted a multi-instrument Mendelian randomization analysis and found that the genetic variation that supports a longer life is significantly associated with the lower risk of COVID-19 infection. The odds ratio is 0.31 (95% CI: 0.18 to 0.52; *P* = 9.7× 10^−6^) per additional 10 years of life, and 0.53 (95% CI: 0.43 to 0.65; *P* = 2.3 × 10^−9^) per unit higher log odds of surviving to the 90th percentile. On the other hand, there was no association between COVID-19 susceptibility and healthspan (the lifespan free of the top seven age-related morbidities). We further applied aging clock models and detected an association between biological age acceleration and future incidence and severity of COVID-19 infection for all subjects as well as for individuals free of chronic disease. Biological age acceleration was also significantly associated with the risk of death in COVID-19 patients. Finally, a bivariate genomic scan for age-related COVID-19 infection identified a key contribution of the Notch signaling pathway. Our analysis suggests that Notch2 expression is associated with a higher risk of COVID-19 infection, providing a druggable target. More generally, interventions that reduce biological age have the opportunity to reduce the risk of COVID-19.

## Introduction

The coronavirus disease 2019 (COVID-19), caused by severe acute respiratory coronavirus 2 (SARS-CoV-2), first emerged in late 2019 and has led to an unprecedented global health crisis^1^. Notably, the aging population is particularly at risk of COVID-19^2^, e.g. in Italy, 88% of the individuals tested positive for COVID-19 were 40 years or older^3^. A recent report based on epidemiological data from multiple countries showed that 69% of infections in people over 70 are symptomatic, whereas this number drops to 21% for 10-19-year-olds^4^. Unsurprisingly, elderly people are also more likely to die from COVID-19, and the case fatality rate for COVID-19 grows exponentially with age^3^. As observational evidence implies a strong link between COVID-19 and age, COVID-19 can be considered a disease of aging^3^, and multiple clinical trials using potential lifespan-extending drugs (e.g., metformin, rapamycin, and senolytics) to protect the elderly from COVID-19 have been proposed^5–7^. Although observational data on metformin seems promising^8,9^, it is unclear if other lifespan-extending drugs should be prioritized in clinical trials, since the evidence of any causal link between lifespan and COVID-19 susceptibility is still missing.

Mendelian Randomization (MR) is a genetic instrumental variable approach that assesses the causal effect of exposure of interest on an outcome, by ascertaining on genetic variants, e.g., single nucleotide polymorphisms (SNPs), strongly associated with the exposure phenotype. Since the alleles of the genetic variants are naturally randomly allocated at conception, when the genetic effects on the outcome are only mediated through the exposure, the causal effect inferred by MR is, in analogy to randomized clinical trials (RCTs), free of any environmental confounders and reverse causation. Although RCTs are considered a gold standard for establishing causal relationships, MR can provide valuable insights into causality when it is not feasible to perform an RCT or before an RCT is performed^10^.

In this study, we performed a multi-SNP MR analysis to elucidate whether and how aging is associated with COVID-19. We considered four lifespan-related traits (parental lifespan, healthspan, longevity, and healthy aging, the combination of these three traits), four measures of epigenetic age acceleration, and four leading genetic risk factors associated with earlier death in humans (Alzheimer Disease (AD), cardiovascular disease (CVD), type 2 diabetes (T2D), and smoking as exposures and evaluated their causal effects on COVID-19 infection and related phenotypes. To support the argument, we also estimated the biological age acceleration (BAA) in COVID-19 patients from UK Biobank (UKBB) cohort and observed a significant association between the phenotypic indicators of aging progress (aging clocks) and the risk and case fatality rate of COVID-19. To provide functional insight into how aging contributes to a higher risk of COVID-19, we further conducted a bivariate genomic scan to highlight the loci contribute to both aging and COVID-19 risk. The pathway enrichment of these loci points to the Notch signaling pathway and specifically to Notch2, whose expression supports an elevated risk of COVID-19 infection.

## Methods

### GWAS data for lifespan-related traits and diseases

We studied four lifespan-related traits with publicly available GWAS summary statistics:

The parental Lifespan GWAS includes unrelated, European-ancestry subjects (a total of 512,047 mother and 500,193 father lifespans), 60% of which were complete. The statistics for every cohort was calculated by fitting Cox survival models to mother and father survival respectively, taking account of 10 principal components, study-specific covariates, and individual sex. Note that in the GWAS setting, parental lifespan is the same phenotype as general lifespan of individuals. This is due to the mathematical property that the genetic effect on a parental phenotype is simply half that on the individual’s phenotype itself. Therefore, the parental lifespan GWAS is a general lifespan GWAS with weaker power. But thanks to the large sample size especially in UK Biobank, such a GWAS is powerful enough to uncover some genetic architecture^11^.

The longevity GWAS includes unrelated, European-ancestry subjects who had a lifespan above the 90th survival percentile (N = 11,262) or whose age at the last follow-up visit (or age at death) was before the 60th percentile age (N = 25,483). The statistics for each cohort were calculated using logistic regression and then combined using a fixed-effect meta-analysis^12^.

The healthspan GWAS contains 300,477 unrelated, British-ancestry individuals from UKBB. The statistics were calculated by fitting Cox-Gompertz survival models, and the events are defined as the first incidence of dementia, congestive heart failure, diabetes, chronic obstructive pulmonary disease, stroke, cancer, myocardial infarction, or demise^13^.

The summary association statistics of healthy aging is from the meta-analysis of healthspan, lifespan, and longevity summary statistics using MANOVA^14^, while accounting for correlations between studies due to sample overlap and correlation amongst the traits. Summary association statistics were calculated for the 7,320,282 SNPs shared between studies. These statistics represent the significance of each SNP affecting one or more of the traits, giving a P-value against the null hypothesis that effect sizes are zero in all studies^14,15^.

We investigated four additional traits genetically correlated with lifespan, using the published case-control studies: Alzheimer’s disease^16^, coronary artery disease^17^, type 2 diabetes^18^, and smoking^19^ (Table S1).

We also included GWAS for age acceleration measured by four epigenetic clocks, including Hannum age, Horvath age, PhenoAge, and GrimAge^20^. The epigenetic age was measured on 34,449 healthy individuals of European ancestry.

GWAS data for 22 common diseases were from a community-based study, Genetic Epidemiology Research on Adult Health and Aging (GERA)^21^, and was analyzed in the original GSMR study^22^. There were 60,586 individuals of European ancestry in the GERA data. There is an additional trait “disease count”, which represents the number of diseases affecting each individual and the summary statistics of these diseases were adjusted with age, gender, and the first 20 PCs.

We used 1000 Genomes Phase 3 reference (released in 2014 October) to map variants in the GWAS results to rsIDs by chromosome, position, and alleles. Only the autosomal SNPs available in the 1000 Genomes reference panel were used, and the 1000 Genomes European ancestry reference was used to estimate the linkage disequilibrium (LD) among these SNPs. Duplicated rsIDs in the data were removed prior to the analysis.

### COVID-19-related traits

To extensively evaluate the genetic effects on COVID-19 risk, we used GWAS summary statistics data from 12 COVID-19-related traits (Table S1). The GWAS results for SARS-COV-2 infection are from the National Institute of Health, Genome-Wide Repository of Associations Between SNPs and Phenotypes (NIH-GRASP), released in August 2020, which includes 1,503 positive cases and 11,409 negative or 457,747 UK Biobank controls with European ancestry. The GWAS summary statistics for severe COVID-19 with respiratory failure is from a genome-wide association study performed in 1,610 cases and 2,205 controls in Italy and Spain^23^. The rest of the 9 traits are from the COVID-19 Host Genetics Initiative (HGI), with the sample size varies from 1,332 to 1,079,768^24^. Among them, 3 are were from HGI release 2 (May 2020), including COVID-19 Hospitalization (versus non-hospitalized COVID-19), susceptibility (affected versus unaffected population), and COVID-19 predicted by flu-like symptoms; Other 6 traits are were in HGI release 3 (June 2020), including very severe respiratory confirmed COVID-19 (versus the general population), COVID-19 infection (versus negative control or population), hospitalized COVID-19 (versus not hospitalized COVID-19 or population), and predicted COVID from self-reported symptoms (versus predicted or self-reported non-COVID-19).

### Expression quantitative trait loci (eQTLs) and age-related gene expression in blood

Blood eQTL data were obtained from the eQTLGen Consortium (31,684 whole blood samples)^25^. Only the significant near-independent eQTLs (FDR-q < 0.05, r^2^ < 0.05) were used in the Mendelian Randomization analysis.

The age-related transcriptomic change in whole blood was obtained from a large-scale meta-analysis^26^, which include six European-ancestry studies (n = 7,074 samples) and detected roughly half of the genes in the human genome (n = 11,908). The direction and P-value of age-related differential expression were directly obtained from the published dataset.

### Mendelian Randomization analysis

Mendelian randomization is a method that uses genetic variants as instrumental variables to determine whether an observational association between a risk factor and an outcome is consistent with a potential causal effect^27^. The multi-SNP MR analysis was implemented using GSMR (Generalized Summary-data-based Mendelian randomization) in GCTA^22^.

As instruments for each exposure (four lifespan-related traits, four risk factors, and four epigenetic age acceleration traits), we selected near-independent SNPs (r^2^ < 0.1) with genome-wide significant (*P* < 5×10^−8^) association with the exposure; For the expression of Notch1-4 in whole blood, we selected significant near-independent eQTLs (FDR-q < 0.05, r^2^ < 0.05); For 22 diseases from GERA community-based study, we selected SNPs with suggestive genome-wide significance (*P* < 1×10^−6^) as instruments and performed a separate analysis due to the limited case number in the community-based study. Full list of the genetic instruments are shown in Supplementary data 1. GSMR includes a HEIDI-outlier filter to remove potential pleiotropic SNPs that have effects on the exposures and the outcomes via different pathways. We set its p-value threshold to 0.01 and tested the remaining SNPs for association with the COVID-19-related traits. The required minimum number of instrumental SNPs for each exposure in the analysis is lowered to 1.

### Bivariate genomic scan and functional Annotation

To identify genetic variants associated with aging-related COVID-19 risk, we meta-analyzed UKBB COVID-19 infection (with population control) and healthy aging (with the sign of effect size reversed) summary statistics, while accounting for correlations between studies due to sample overlap and correlation between the traits, as implemented in MultiABEL v1.1-610^14,28^, Summary association statistics were calculated for the 7,318,649 SNPs shared between studies. These statistics represent the significance and consistency of each SNP affecting one or both of the traits (e.g. the SNPs that significantly contribute to both aging and COVID-19 risk in the same direction will have a smaller P-value). Therefore, we refer to this bivariate genomic scan result as the aging-related COVID-19 throughout this study.

We then used the summary statistics of aging-related COVID-19 and performed functional annotation for all SNPs in genomic areas identified by lead SNPs (*P* < 1 × 10^−6^, 250 Kb apart) using FUMA (Functional Mapping and Annotation)^29^. The annotated genes were then used for functional enrichment analysis using the default setting on the FUMA platform.

### Genetic correlation analysis

We estimated the genetic correlations between lifespan-related traits, risk factors, epigenetic age acceleration, and COVID-19 using LD score regression (LDSC) and high-definition likelihood (HDL) methods^30,31^, SNPs that are imperfectly imputed (INFO < 0.9) or with low frequency (MAF < 0.05) were removed to reduce statistical noise. LDSC was performed using LDSC software v1.0.1 (https://github.com/bulik/ldsc); and the HDL was performed using R package “HDL” v1.3.8 (https://github.com/zhenin/HDL).

### Biological age estimation for UKBB cohorts

All-cause mortality increases exponentially with age, and hence log-linear risk predictors from proportional hazards models can provide natural composite organism state representations characterizing the progress of aging based on biological and physiological measurements. We used two such biological age measures: Phenotypic Age based on blood biochemistry^32^, and Dynamic Organism State Indicator (DOSI) based on widely available Complete Blood Counts (CBC) only^33^. The latter is a proxy for the frailty index and is derived from the blood markers only, whereas the Phenotypic Age additionally employs the explicit age. We also used physical activity (the number of steps per day averaged over the week), which is also associated with all-cause mortality and hence can also be viewed as a measure of biological aging^34^.

We investigated the association of the incidence of COVID-19 with biological aging acceleration (BAA, which is the difference between the biological age of a person and the average biological age in the cohort of individuals of the same age and sex) using logistic regression. Chronological age and biological sex were used as additional covariates in the analysis.

Following UKBB recommendations, we used the “result” label from the table “COVID-19 test results table” as the proxy of disease severity. This implies that mostly those individuals that showed characteristic COVID-19 symptoms were selected for testing. We investigated BAA associations with the incidence of COVID-19 and its associated fatality using all available cases (All) and separately cohorts of individuals who have (Frail) or do not have (Not Frail) major chronic diseases (from the list including all kinds of cancer, angina pectoris, coronary heart disease, heart attack, heart failure, hypertension, stroke, diabetes, arthritis, bronchitis, and emphysema) at the time of infection.

## Results

We applied GSMR to test for potential causal associations between four lifespan-related traits and COVID-19, including lifespan, longevity (i.e. surviving to the 90th percentile), healthspan (time to a first major age-related disease), and healthy aging (multivariate meta-analysis of all three traits combined) (Table S1). We employed summary-level GWAS data^11–13,15^ and selected near-independent SNPs at a genome-wide significance level as genetic instruments for each trait. The HEIDI-outlier filter was used to detect and eliminate genetic instruments with pleiotropic effects on both exposure and outcome, as described by Zhu et al.^22^. For the outcomes, we used 12 different sets of GWAS summary statistic data for COVID-19-related traits from case-control studies (Table S1).

Strikingly, our GSMR analysis showed that long lifespan, longevity, as well as healthy aging were protective against COVID-19 infection based on UKBB reporting (Fig. 1 A-E, Table 1). The estimated odds ratio for lifespan was 0.31 (95% CI: 0.18 to 0.52; *P* = 9.7 × 10^−6^), indicating that the risk of COVID-19 infection is decreased by 69% with approximately every additional 10 years of life^11^. For longevity, the odds ratio was 0.53 (95% CI: 0.43 to 0.65; *P* = 2.3 × 10^−9^), and for healthy aging, the odds ratio was 0.12 (95% CI: 0.05 to 0.32; *P* = 1.6 × 10^−5^). As UKBB reporting on COVID-19 infection is biased toward severe cases of hospitalized subjects, this measure might also be related to COVID-19 severity and mortality^35^. We observed a similar effect of these three traits on COVID-19 susceptibility in the HGI dataset with overlapping confidential interval of estimated effect size, suggesting the association between lifespan-related traits and COVID-19 infection is robust across different cohorts (Fig. 1A, Table 1). However, none of the lifespan-related traits showed a significant protective effect on COVID-19 with a severe respiratory disorder or respiratory failure, possibly due to the small case number of severe COVID-19 (Table S1). We also estimated the causal effect of genetically proxied epigenetic age acceleration on the risk of COVID-19^20^, while none of them shown to have a significant effect on COVID-19 after Bonferroni correction for 144 tests (Fig. S1).

**Table 1.** Mendelian randomization estimates for the association between lifespan-related traits and risk of COVID-19.

| *Exposure* | *Outcome* | *OR* | *95% CI* | *P* |
| --- | --- | --- | --- | --- |
| *Healthy aging* | HGI_covid_susceptibility | 0.33 | 0.13 - 0.85 | 2.2e-02 |
|  | UKBB_covid_vs_neg | 0.25 | 0.09 - 0.69 | 7.0e-03 |
|  | UKBB_covid_vs_pop | 0.12 | 0.05 - 0.32 | 1.6e-05 |
| *Lifespan* | HGI_covid_susceptibility | 0.45 | 0.27 - 0.77 | 3.2e-03 |
|  | HGI_covid_vs_neg | 0.61 | 0.39 - 0.97 | 3.5e-02 |
|  | UKBB_covid_vs_neg | 0.44 | 0.26 - 0.77 | 3.6e-03 |
|  | UKBB_covid_vs_pop | 0.31 | 0.18 - 0.52 | 9.7e-06 |
| *Longevity* | HGI_covid_susceptibility | 0.68 | 0.56 - 0.82 | 8.5e-05 |
|  | UKBB_covid_vs_neg | 0.58 | 0.47 - 0.72 | 5.0e-07 |
|  | UKBB_covid_vs_pop | 0.53 | 0.43 - 0.65 | 2.3e-09 |

Only the associations that reached nominal significance ( $P < 0.05$ ) are shown

**Figure 1.**
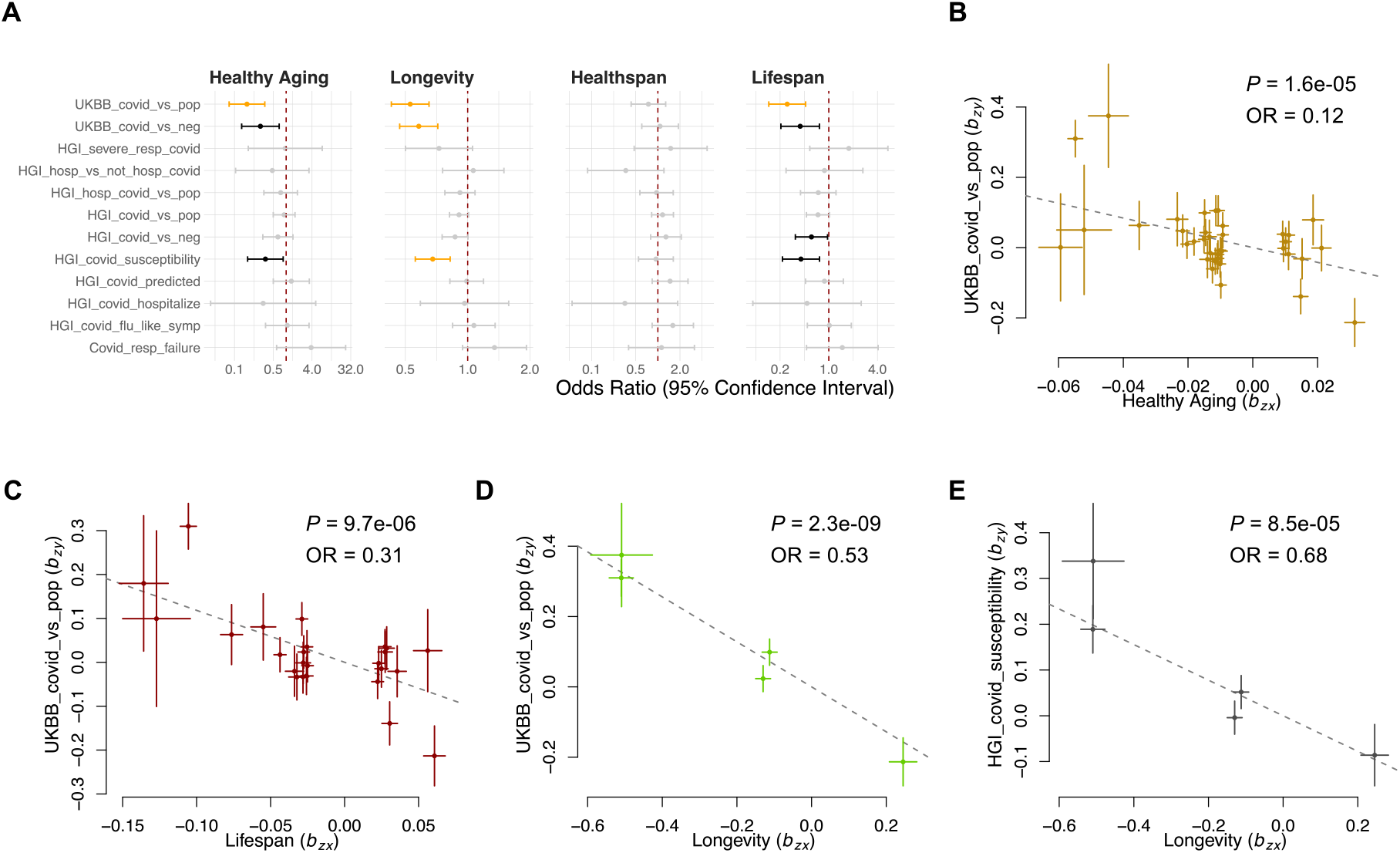
Mendelian Randomization analysis investigating the association of lifespan-related traits with the risk of COVID-19. **A.** The forest plot showing Mendelian randomization estimates for the causal effect of lifespan-related traits on the risk of COVID-19. Error bars show the 95% confidential interval. Significant effects after correcting for 144 tests (12 exposures and 12 outcomes, *P* < 0.05/144) are in orange. Nominally significant effects (*P* < 0.05) are in black. **B-E**. Plots of effect sizes of all genetic instruments from GWAS for healthy aging **(B)**, lifespan **(C)**, and longevity **(D)** (x-axis) versus those for UKBB COVID-19 (y-axis); and longevity (x-axis) versus HGI COVID-19 (y-axis) **(E)**. Error bars represent standard errors.

The loci for AD, CVD, T2D, cancer, and smoking (or lung cancer) explained the most genetic variance of lifespan, as reported by Timmer et al.^11^. To investigate whether these risk factors contribute to the plausible causal association between lifespan and COVID-19, we conducted an MR analysis of late-onset AD, coronary artery disease (CAD, including myocardial infarction, percutaneous transluminal coronary angioplasty, coronary artery bypass grafting, chronic ischemic heart disease, and angina), T2D, and smoking (the number of cigarettes smoked per day) as exposures (Fig. S1 A-C, Supplementary data 2). Only the late-onset AD was found to significantly increase the risk of COVID-19 after Bonferroni correction for 144 tests (*P* = 4.3×10^−6^), with the odds ratio of 1.14 (95% CI: 1.08 to 1.21), suggesting the benefit of a longer lifespan on the risk of COVID-19 may be partially mediated by less severe or later occurring Alzheimer’s disease.

To further evaluate the risk factors for COVID-19 infection and severity, we conducted a separate MR analysis using GWAS data of 22 common diseases from GERA^21^ (Fig. S3, Table S3). None of the diseases reached the significance threshold after Bonferroni correction (*P* = 0.05/276 = 2 × 10^−4^). Among the nominally significant associations, asthma, dyslipidemia, hernia abdominopelvic cavity, peptic ulcer, hypertensive disease, age-related macular degeneration, and allergic rhinitis were the risk factors for COVID-19 infection; hypertensive disease, irritable bowel syndrome, peripheral vascular disease, age-related macular degeneration, and varicose veins were the risk factors for COVID-19 severity (Fig. S3, Supplementary data 2). Interestingly, the disease count, a trait that represents the number of comorbidities affecting each individual, was the risk factor for severe cases of COVID-19 (hospitalization). The odds ratio was 3.57 (95% CI: 1.04 to 12.28, *P* = 0.04), suggesting a subject with roughly every two (one standard deviation) more comorbidities has a 3.6-fold higher risk of having a severe case of COVID-19 (Table S2).

Healthspan is defined as the age period free of major age-related morbidities. In the healthspan GWAS study, the top seven age-related morbidities were included (see Method)^13^. In our analysis, healthspan did not show a significant effect on COVID-19-related traits (Fig 1A). This is unlikely to be due to the power of healthspan GWAS since there were 17 near-independent genome-wide significant SNPs (*P* < 5 × 10^−8^), which is more than lifespan and longevity. Therefore, we hypothesized that the strong protective effect of longevity against COVID-19 may not be explained by the delayed appearance of age-related morbidities, but rather by decelerated biological age that extends lifespan.

To address this hypothesis, we assessed in parallel the three different risk-based biological age predictions computed for the subjects in the UKBB cohort using blood biochemistry (Phenotypic Age), Complete Blood Counts (DOSI), and physical activity measurements^32–34^ (Fig. 2A). We found that COVID-19 incidence in all UKBB datasets was significantly associated with the BAA of Phenotypic Age, DOSI, and decreased physical activity (Fig. 2B-E, Table 2). The estimated odds ratio of COVID-19 infection is 1.28 (95% CI: 1.25 to 1.31; *P* = 8.4 × 10^−82^) and 1.31 (95% CI: 1.26 to 1.38; *P* = 9.5 × 10^−32^) for every ten years higher biological age measured by Phenotypic Age and DOSI, respectively. Phenotypic Age and DOSI were also significantly associated with COVID-19 incidence and case fatality independently from the BAA association with chronic diseases, i.e., separately in cohorts of UKBB individuals having (Frail) or not (Not frail) chronic age-related health conditions (Fig. 2E, Table 2).

**Table 2.**
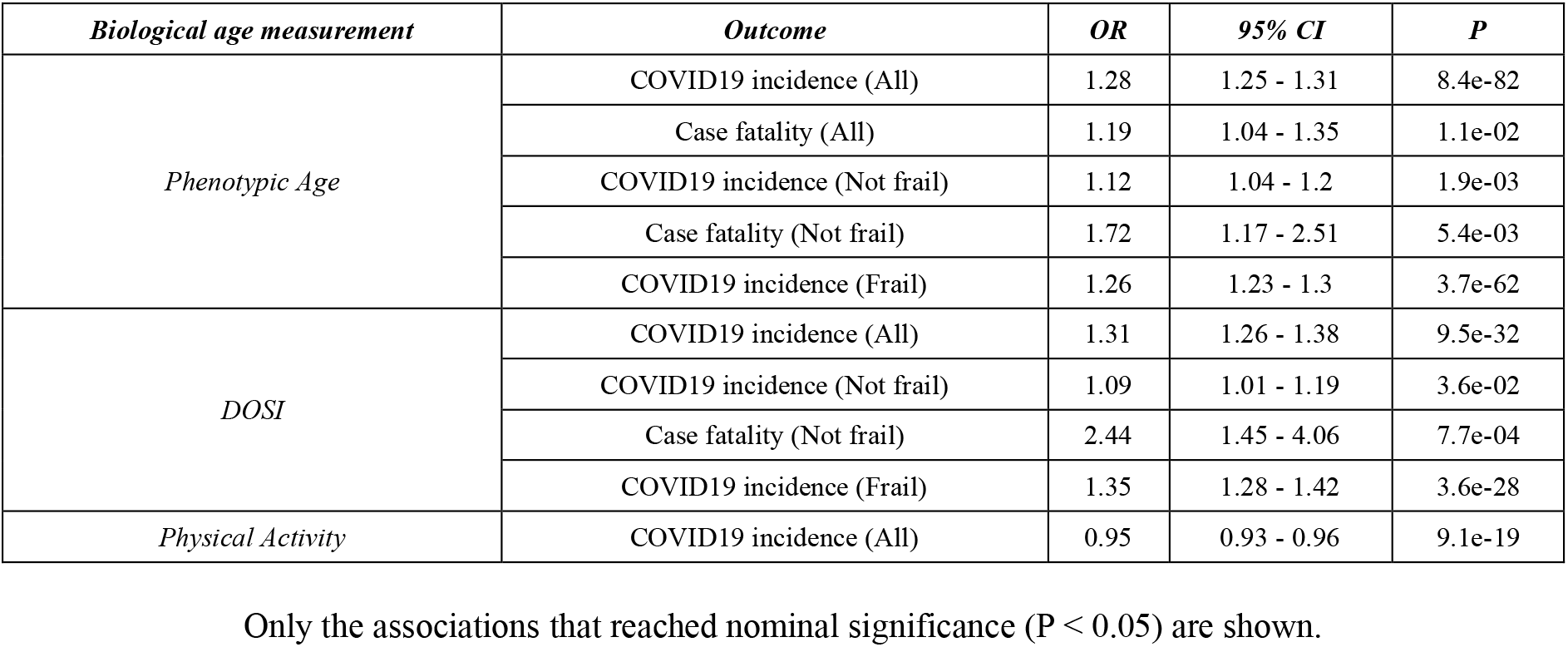
Association between biological age acceleration and the risk of COVID-19.

| *Biological age measurement* | *Outcome* | *OR* | *95% CI* | *P* |
| --- | --- | --- | --- | --- |
| *Phenotypic Age* | COVID19 incidence (All) | 1.28 | 1.25 - 1.31 | 8.4e-82 |
|  | Case fatality (All) | 1.19 | 1.04 - 1.35 | 1.1e-02 |
|  | COVID19 incidence (Not frail) | 1.12 | 1.04 - 1.2 | 1.9e-03 |
|  | Case fatality (Not frail) | 1.72 | 1.17 - 2.51 | 5.4e-03 |
|  | COVID19 incidence (Frail) | 1.26 | 1.23 - 1.3 | 3.7e-62 |
| *DOSI* | COVID19 incidence (All) | 1.31 | 1.26 - 1.38 | 9.5e-32 |
|  | COVID19 incidence (Not frail) | 1.09 | 1.01 - 1.19 | 3.6e-02 |
|  | Case fatality (Not frail) | 2.44 | 1.45 - 4.06 | 7.7e-04 |
|  | COVID19 incidence (Frail) | 1.35 | 1.28 - 1.42 | 3.6e-28 |
| *Physical Activity* | COVID19 incidence (All) | 0.95 | 0.93 - 0.96 | 9.1e-19 |

Only the associations that reached nominal significance ( $P < 0.05$ ) are shown.

**Table 3.** Mendelian randomization estimates for the association between Notch2 expression and risk of COVID-19.

| *Exposure* | *Outcome* | *OR* | *95% CI* | *P* |
| --- | --- | --- | --- | --- |
| *NOTCH2* | HGI_covid_vs_neg | 1.30 | 1.03 - 1.65 | 0.03020 |
|  | HGI_covid_vs_pop | 1.31 | 1.1 - 1.55 | 0.00197 |
|  | UKBB_covid_vs_neg | 1.46 | 1.08 - 1.99 | 0.01540 |
|  | UKBB_covid_vs_pop | 1.43 | 1.07 - 1.91 | 0.01460 |

Only the associations that reached nominal significance ( $P < 0.05$ ) are shown.

**Figure 2.**
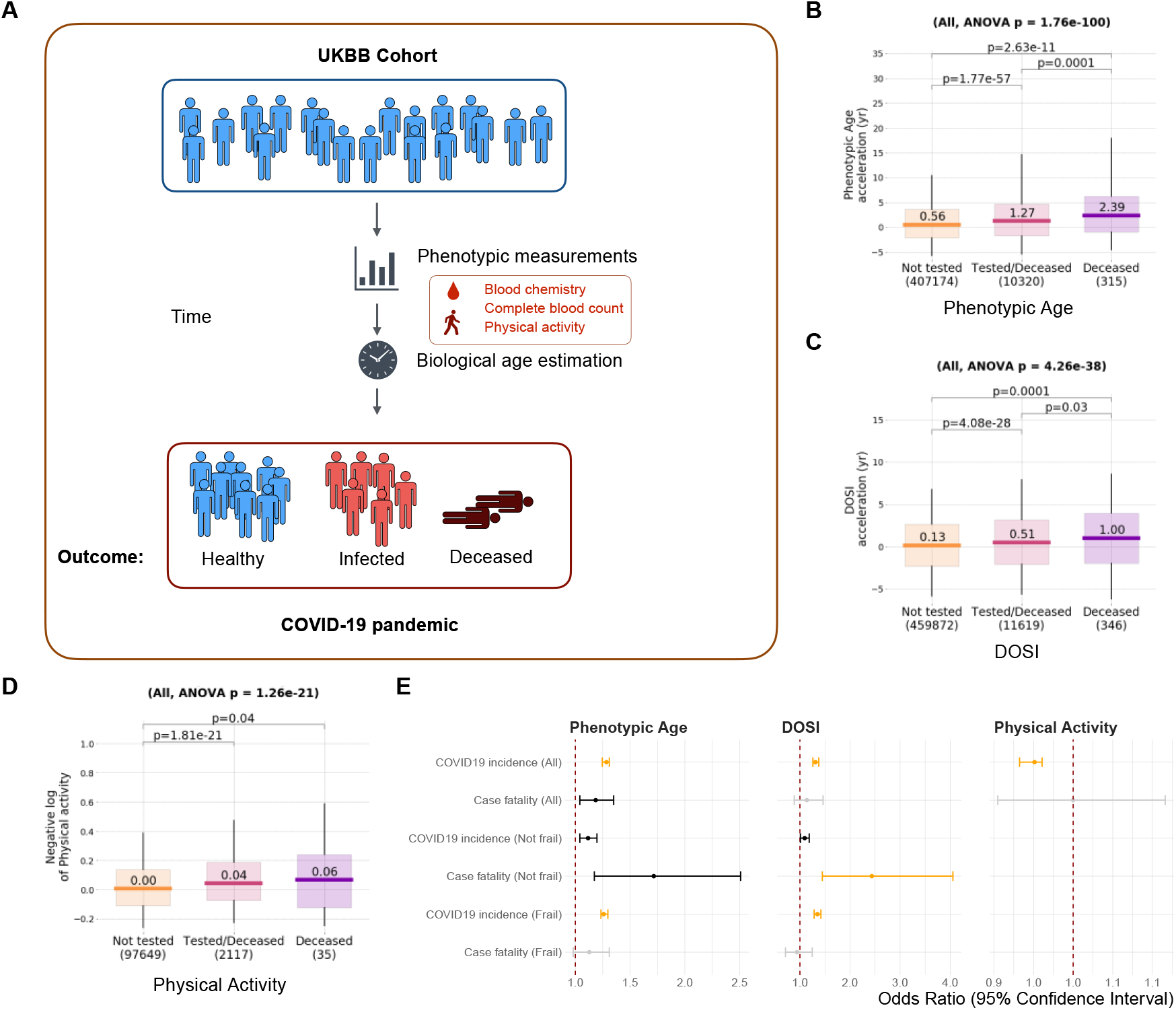
Analysis of association of biological age acceleration with the risk of COVID-19. **A.** Schematic representation of analysis of the biological age acceleration in UKBB cohort. **B-D**. the box plots showing the distribution of biological age acceleration measured by phenotypic age **(B)**, DOSI **(C)**, and negative log physical activity **(D)** in UKBB subjects that were not tested, tested, or died with COVID-19 infection. **E**. The forest plot showing the predicted effect of biological age acceleration on the risk of COVID-19 in different catagory. Error bars show the 95% confidential interval. Significant effects (P < 0.001) are in orange. Nominally significant effects (P < 0.05) are in black. Odds ratio for Phenotypic Age and Dynamic Organism State Index (DOSI) is given per 10-yr biological age acceleration. Odds ratio for physical activity is given per increase of 1000 steps/day.

We also observed elevated BAA levels of all measures of biological age (Fig. 2B-D, Fig. S2A-D) in cohorts of individuals died from COVID-19 compared to those tested (and most probably suffering from the disease), and, separately, in cohorts of those tested versus the rest of UKBB (and presumed free of the disease). The number of UKBB subjects with data fields required for the Phenotypic Age and DOSI was comparable, and we found that Phenotypic Age comparisons produced a better statistical power. The number of UKBB subjects with physical activity metrics was small, but the association of BAA in the form of physical activity deficit and the incidence of COVID-19 was significant.

To gain the mechanistic insight of how aging and COVID-19 intertwined at the genetic level, we performed a bivariate genomic scan using the GWAS of healthy aging and UKBB COVID-19 infection, to identify the genetic variants that contribute to both aging and the risk COVID-19 infection i.e., aging-related COVID-19 risk (Fig. S4, see Method). We identified twenty bivariate loci at genome-wide significance (*P* < 5 × 10^−8^), where the null hypothesis is no association with healthy aging and COVID-19 infection (Fig. S4). The summary statistics of aging-related COVID-19 risk were then annotated using FUMA and a functional enrichment analysis in 2868 canonical pathways (including gene sets from BIOCARTA, KEGG, PID, REACTOME, and WikiPathways) and 7350 Gene Ontology (GO) biological processes was performed. We find significant enrichment (P_adjusted_ < 0.05) in 67 canonical pathways and 26 biological processes. The canonical pathways with the strongest enrichment include pre-Notch expression and processing (*P* = 3.0 × 10^−8^), signaling by Notch (*P* = 3.6 × 10^−7^), and oxidative stress-induced senescence (*P* = 1.4 × 10^−6^) (Fig. 3A, Supplementary Data 3). Top enriched biological processes are immune system development (*P* = 2.3 × 10^−7^) and myeloid cell differentiation (*P* = 2.4 × 10^−6^), among others (Fig. 3B, Supplementary Data 3).

**Figure 3.**
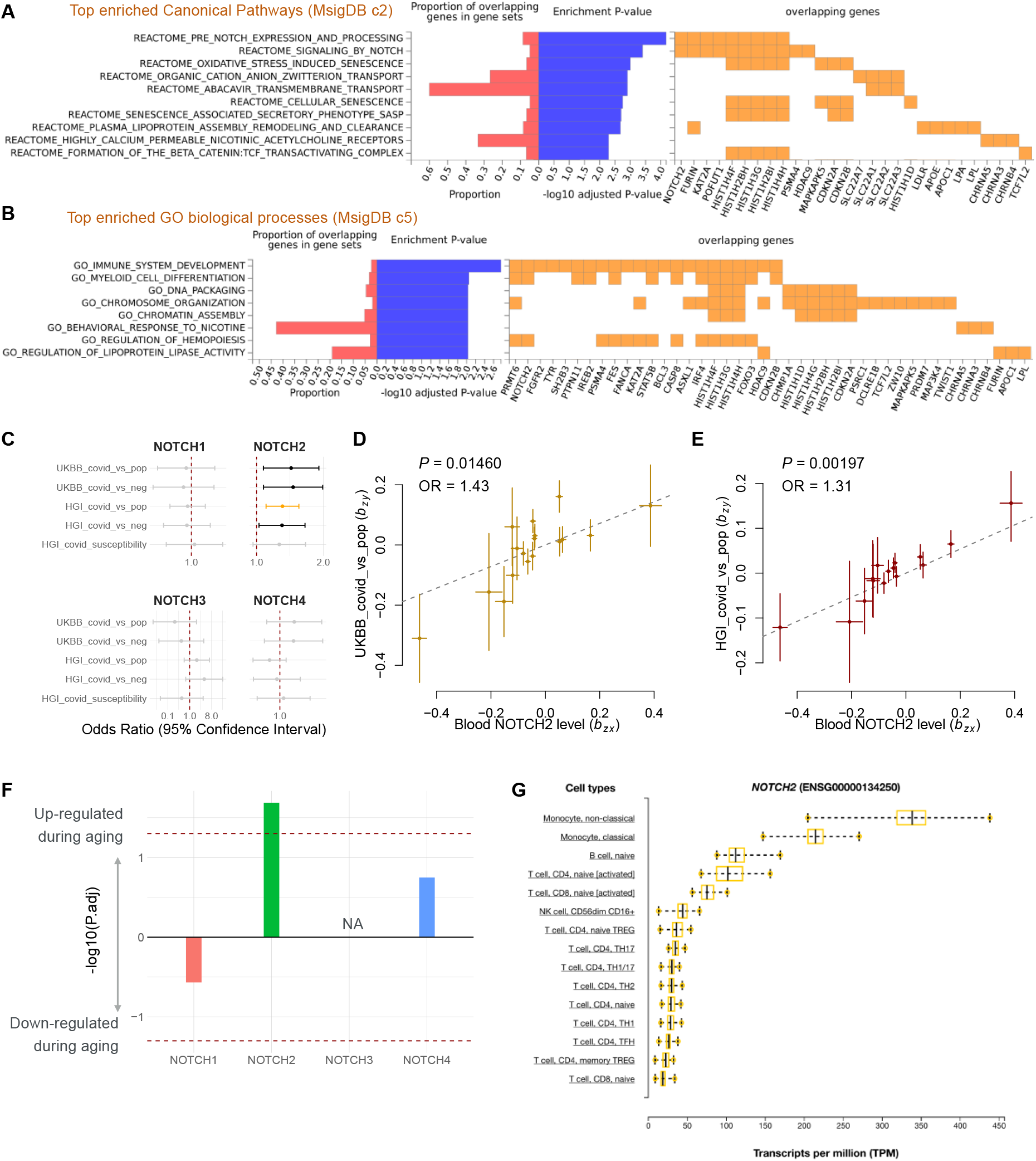
Bivariate genomic scan identifying Notch signaling as age-related COVID-19 risk. **A, B**. Gene set enrichment analysis of aging-related COVID-19. Top significantly enriched (P_adjusted_ < 0.05) canonical pathways **(A)** and GO biological processes **(B)** are shown. **C**. Forest plot showing Mendelian randomization estimates for the causal effect of blood Notch2 expression on the risk of COVID-19 infection. Error bars show the 95% confidential interval. Significant effects after correcting for 20 tests (*P* < 0.0025) are in orange. Nominally significant effects (*P* < 0.05) are in black. Error bars show the 95% confidential interval. **D, E**. Plots of effect sizes of all genetic instruments from blood eQTLs for Notch2 (x-axis) versus those for UKBB **(D)** and HGI **(E)** COVID-19 infection (y-axis). Error bars represent standard errors. **F**. Bar plot showing the age-related differential expression of Notch1-4 in blood. The y-axis represents the −log10(P.adj) × sign of changing direction, i.e. positve value represents an age-related increase. **G**. Expression levels of Notch2 in transcripts per million (TPM, x-axis) from the DICE database. Cell types (y-axis) are sorted based on the median expression level within the cohort from highest to lowest. Boxes indicate 25%-75% interquartile ranges, and whiskers indicate minimum to maximum.

The Notch pathway is an evolutionally conserved signaling pathway, which is suggested to be involved in both age-related inflammation and the development of age-related disease^36^. Moreover, the Notch signaling is related to the entry of SARS-CoV-2 through the positive regulation of the host proteins that promote the entrance of the virus into the cell (e.g. FURIN and ACE2)^37^. In humans, there are four paralogs in the Notch family (Notch1-4)^38^. We hypothesized that the Notch signaling is a mediator for aging-related COVID-19 infection and its effect may be related to the expression of Notch. This hypothesis was investigated with MR of blood eQTLs of Notch1-4 from eQTLgen^39^, against five traits that representing COVID-19 infection. We found that the expression of Notch2 significantly (*P* < 0.05/20) increases the risk of COVID-19 infection (Fig. 3 B-D). The odds ratio estimated from HGI COVID-19 GWAS was 1.31 (95% CI: 1.1 to 1.55; *P* = 0.002) per standard deviation higher Notch2 expression in blood. We also observed a similar odds ratio estimate with overlapping 95% CI in other four COVID-19 infection traits. This result suggests a causal role of Notch2 and more generally Notch signaling in COVID-19 infection. To determine the age-related expression of Notch, we examined the dataset of Peters et al.^26^, which contains associations of genes with age in humans, estimated from 7,074 whole blood samples. Among Notch1-4, only the Notch2 significantly (*P* = 0.007) increased during aging, suggesting that the age-related increase of COVID-19 risk is partially mediated through the increase of Notch2 expression. Moreover, we found that Notch2 is primarily expressed in monocytes based on DICE (database of immune cell expression, eQTLs, and epigenomics) project (Fig. S5)^40^, which is also implicated in aging and COVID-19^41^. This finding offers a new druggable target for preventing COVID-19 in the elderly and is worthy of further experimental studies.

Finally, we estimated the genetic correlations between lifespan-related traits and COVID-19 using LD score regression and high-definition likelihood (HDL) methods (Fig. 4, supplementary data 4)^30,31^, However, among 12 COVID-19-related traits, we only observed the the positive genetic correlation between COVID-19 infection and AD at nominal significance level, suggesting the case sample sizes of COVID-19 GWAS studies were too small to support a sufficient power for estimating genetic correlations among these traits. Future genetic analyses utilizing larger sample sizes should provide opportunities to improve these estimates.

**Figure 4.**
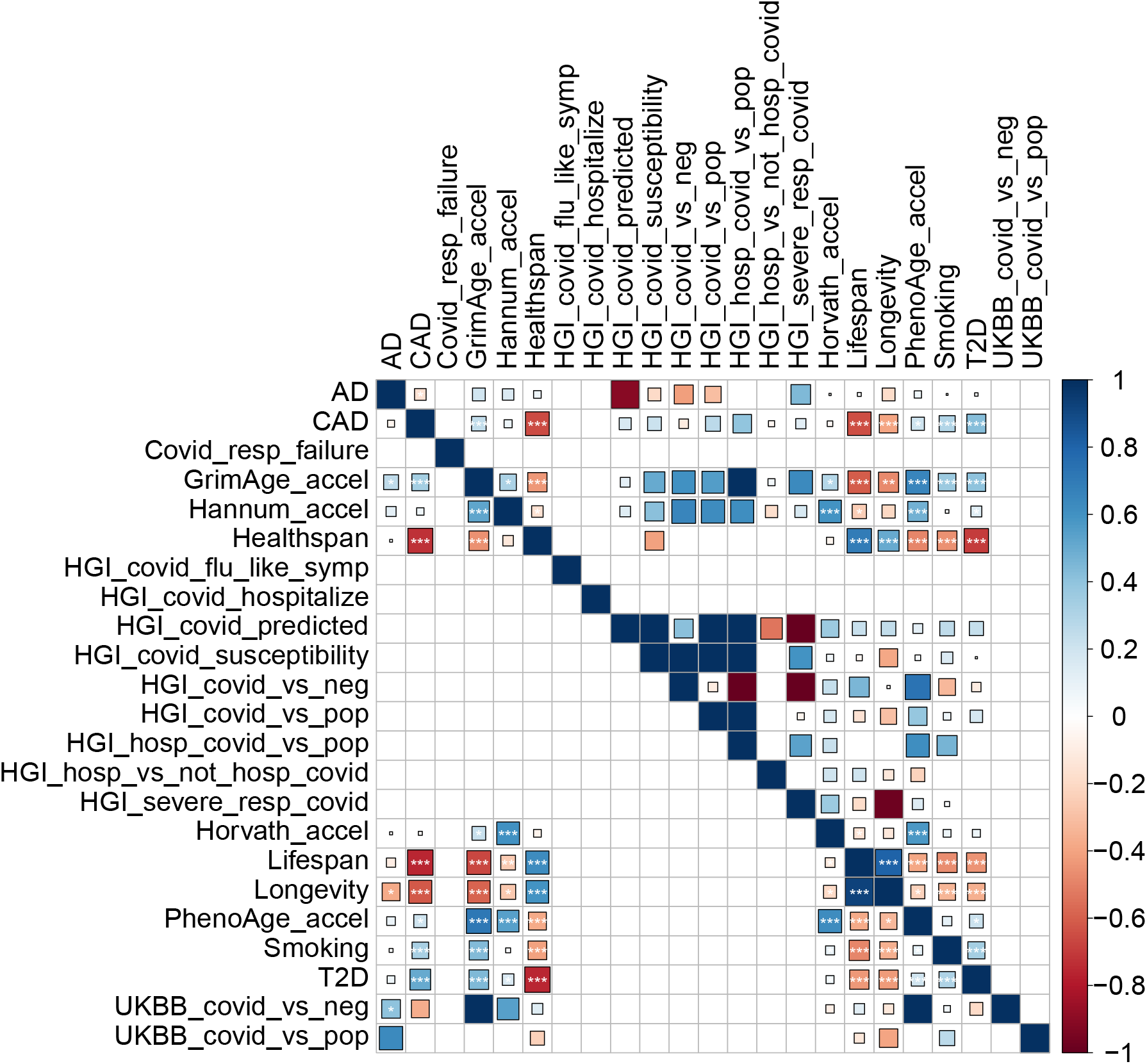
Genetic correlation estimates from HDL and LDSC among phenotypes. Lower triangle: HDL estimates; upper triangle: LDSC estimate. the areas of the squares represent the absolute value of corresponding genetic correlations. The genetic correlation that couldn’t be estimated are in blank. P values are corrected using Bonferroni correction for 253 tests, * P_nominal_ < 0.05, ** P_adjusted_ < 0.05, *** P_adjusted_ < 0.01.

## Discussion

In this study, we explored a potential causal relationship between aging and the risk of COVID-19 by conducting a multi-instrument MR analysis using four different lifespan-related traits as exposures and eleven COVID-19-related traits as outcomes. We found that genetically proxied longer lifespan and longevity were significantly associated with the decreased risk of COVID-19 (*P* = 9.7× 10^−6^ and 2.3 × 10^−9^, respectively), and further analyses revealed a key role of an elevated biological age and severity of chronic age-related diseases in this association. One of the contributing factors is likely the immune response. The competence of the immune system declines as people age, which is known as “immunosenescence”^42^. The hallmarks of immunosenescence include an impaired response to new antigens, unsustained memory responses, increased autoimmune responses, and prolonged inflammation. As a result, elderly subjects are more susceptible to infectious diseases, including COVID-19, and have a poor response to vaccines^42,43^. On the other hand, it has been reported that the circulating immune cells in centenarians possess unique characteristics that sustain immune responses to infections^44^. Moreover, the offspring of centenarians were shown to have a lower level of inflammation^45^, suggesting that the benefits on the immune system in centenarians are heritable. Therefore, a better immunological profile in people with pro-longevity genetics may help support the causal effect of longevity on COVID-19 we observed.

Notch pathway is an evolutionally conserved signaling pathway involved in age-related inflammation and disease^36^. Moreover, Notch signaling is related to the entry of SARS-CoV-2 through the positive regulation of host proteins that promote entrance of the virus into the cell^37^. The entry of SARS-CoV-2 is mediated by binding of its S (spike) glycoprotein to the Angiotensin Converting Enzyme 2 (ACE2)^46^. Therefore, upregulation of ACE2 could potentially increase the risk of viral infection. ADAM17 (A Disintegrin And Metalloproteases 17) is a metalloprotease that supports the shedding of ACE2 on the cell membrane^47^. It is negatively regulated by Notch signaling, whereas downregulation of ADAM17 significantly reduced the shedding of ACE2^37^. Besides ADAM17, a proteolytic cut of the S protein mediated by furin after S glycoprotein binds to ACE2 is also required for the entry of SARS-CoV-2 into the cell. The expression of furin is positively regulated by Notch signaling, and furin is also involved in the maturation of Notch precursor^37^. This evidence is in line with our finding that Notch signaling plays an important role in aging-related COVID-19.

Notch2 is one of the four Notch paralogs in mammals. In our MR analysis, we found evidence of the causal relationship between Notch2 expression and COVID-19 infection (Fig. 3 B-D). A previous study suggests that Notch2 promotes goblet cell metaplasia in the lung, which is the hallmark of airway diseases^48^. Moreover, the goblet cells are the major source of ACE2 in the lung and were indicated to play a significant role in enabling COVID-19 infections. Therefore, the increased Notch2 expression during aging might play a causal role in the increased risk of COVID-19 infection in the elderly. We observed a relatively large effect size (31% increased risk of infection for every 1 standard deviation higher Notch2 expression), suggesting the Notch2 might provide a desirable target for the prevention and treatment of COVID-19, as well as identifying population with a higher potential risk of infection. Further experimental and clinical study on Notch2 and COVID-19 is needed to validate the causal relationship.

Aging manifests as progressive remodeling of the organism, and hence a great number of biological measurements are associated with age. Several sets of physiological and biological indices have been proposed for quantification of aging progression in the form of a single number – the biological age^49,50,^ or frailty index^51,52^. One popular approach is to regress relevant variables to predict chronological age and thus produce the “biological age” prediction. Popular Hannum’s and Horvath’s methylation age-clock models, as well as other clocks, are the widely used examples of such an approach^53,54^,

An interesting alternative is to produce the log-linear all-cause mortality estimate with a proportional hazard model and treat the resulting value as a measure of biological age. Phenotypic Age from blood biochemistry markers^32^, DOSI from CBC^33^, averaged physical activity levels^34^, and more sophisticated machine learning algorithms used to predict the risk of death from physical activity time series of wearable devices^55^, or even self-reported health questionnaires, are all examples of this approach^56^. All reliable biological age predictors are associated with chronic disease burden, unhealthy lifestyles such as smoking (both overall and in disease-free population), and future incidence of chronic diseases in healthy subjects^32–34,49,50,52,57,58^. In our work, we also established the association of BAA with the risk of non-chronic diseases, such as COVID-19 and the corresponding case fatality in the UKBB cohort independent of disease burden. The association was significant for BAA measures obtained from blood biochemistry (Phenotypic Age)^32^, CBC (DOSI)^33^ and mean physical activity (number of steps per day recorded by wearable devices over a week-long period of time^34^; the number of UKBB subjects with physical activity measurements was too low for separate BAA characterization in frail and non-frail cohorts).

Decreased physical activity was associated with an increased risk of COVID-19 in the UKBB cohort. This observation may be interesting on its own since the widespread lockdown measures brought about a dramatic (up to 27.3%, which is 1,432 steps per day, within 30 days) decline of average physical activity^59^. Our association study suggests a more than 10% risk increase corresponding to 1.5 thousand steps per day loss. There are feedback loop effects of decreased mobility on BAA measures, and as such, the associated risk adjustments must be taken into account in advanced epidemiological models of lockdown effects.

One advantage of our study design is that all of the BAA predictors were measured prior to the pandemic. Therefore, the association between BAA and the risk of COVID-19 (and probably other dangerous infectious diseases) is free of reverse causation (like in MR) and likely to be causal if there are no other confounders. Thus, our research supports the idea of the pro-active application of anti-aging (that is BAA-reducing) drugs in a prophylaxis mode to protect the biomarker-defined vulnerable individuals. And, reversely, a significant reduction of BA by an experimental drug in a clinical trial (probably as early as phase I) could warrant further clinical studies in elderly subjects.

The association of BAA with case fatality was weaker (only Phenotypic Age BAA exhibited a significant effect). This can be explained by the considerably smaller number of UKBB subjects involved in the statistical analysis (346 of dead individuals compared to 11,619 tested (and presumed sick) and 459,872 overall subjects in UKBB). The case fatality rate increases exponentially with age, and therefore it would be reasonable to expect the association of BAA with the risk of death in COVID-19 patients^3^. We expect future studies to corroborate our findings. Whether or not this association is causative could not be established in our study. The age-dependent severity of COVID-19 has been demonstrated in many epidemiological studies^3,60^, However, we did not observe a significant association between longevity and COVID-19 severity-related traits. This could be largely due to a small number of cases included in GWAS analyses, varying from 536 to 1,610. Moreover, there is an underlying selection bias toward symptomatic cases in the GWAS of COVID-19 positive cases, especially in the UKBB study, which is overrepresented by severe and hospitalized cases^35^. Therefore, our results on UKBB may also be interpreted as a protective effect of longevity on the severe form of COVID-19 infection.

It has been reported that people with comorbidities are more likely to suffer from COVID-19 and have poor prognosis^61^. The most prevalent comorbidities were hypertension, diabetes, and cardiovascular disease^3,61^, Based on our Mendelian randomization analysis, genetically predicted CAD, one of the most common types of cardiovascular disease, increases COVID-19 susceptibility and the chance of being hospitalized after infection at a nominal significance level, which is consistent with the result of recent study^62^. However, we didn’t find evidence of the causal effect of T2D and smoking on COVID-19-related traits. There are two possible explanations: (1) the observed link between T2D and COVID-19 in epidemiological studies is confounded by other factors; (2) the power of current COVID-19 GWAS results is limited to reveal its potential causal link to T2D/smoking. Notably, a nominally significant association (P = 0.03) between the life-time smoking index and risk of severe COVID-19 was reported^63^, suggesting a potential causal role of smoking on COVID-19. Future genetic analyses with larger sample sizes or clinical experiments are required to fully identify the relationship between smoking and COVID-19. The genetically predicted late-onset AD was also shown to be significantly associated with a higher risk of infection based on both UKBB and HGI report, which is not observed in another recent MR analysis^64^. This is possibly due to the larger case number (35,274 compared with 17,008) was included in late-onset AD GWAS, thus increased the power of the MR analysis.

There are multiple clinical trials proposed to employ potential lifespan-extending drugs to protect the elderly from COVID-19, based on promising observational data on metformin^5–9^. However, epidemiological studies are prone to confounding, reverse causation, and various biases, and therefore are an unreliable indicator of the causal associations. MR is a method that utilizes genetic instruments that are robustly associated with exposures, and thus generate more reliable evidence in predicting novel interventions^65^. In our MR analysis, we found evidence for the causal relationship between longevity and decreased COVID-19. The following analysis of genetic risk factors and phenotypic measurements suggests that this causal effect is likely to be mediated by the decelerated rate of aging, which can be captured by biological age measurements. Therefore, our finding can directly support lifespan-extending drugs as a potential measure of COVID-19 when one of the following assumptions holds: (1) the selected anti-aging drugs extend lifespan through a mechanism that mimics the genetics of longevity; and (2) the selected anti-aging drugs could slow down or reverse the aging process measured by biological age models (e.g., phenotypic age).

While the first assumption is hard to test, recent studies suggest that some anti-aging interventions can slow down and even reverse the biological age measured by biological age models^66^. For example, a cocktail treatment of recombinant human growth hormone, dehydroepiandrosterone, and metformin reversed the immunosenescent trend, and the biological age measured by several biological age models (including PhenoAge) was reversed by 2.5 years on average after 12 months of treatment^66^. Thus, it could be worthwhile prioritizing established anti-aging drugs in COVID clinical trials. This may accelerate COVID drug development and save costs.

## Data Availability

This research has been conducted using publicly available GWAS summary statistics (for URLs, see Table S1) and the UK Biobank Resource under Application Number 21988

## Acknowledgements

XS was in receipt of a Swedish Research Council (Vetenskapsrådet) Starting Grant (No. 2017-02543). This research was also supported by NIA grants (to VNG). PF and TP were supported by Gero PTE LLC (Singapore). We thank HGI and NIH-GRASP for the timely release of COVID-19 GWAS summary statistics. We also thank Miss. Hanna Liu for assistance in part of the analysis.

## Author contributions

XS initiated the study; VNG, XS, and POF supervised the study; KY, RZ, and TVP performed data analyses; KY, VNG, POF, and XS wrote the manuscript; All authors contributed to manuscript writing.

## Data availability statement

GWAS summary statistics used in this study are publicly available (for URLs, see Table S1). The individual-level phenotype data are available by application from the UK Biobank (http://www.ukbiobank.ac.uk/). The bivariate GWAS summary statistics of aging-related COVID-19 generated in this study are available at https://www.dropbox.com/s/bt6mtmttzgnhtfo/combined_ukbbCOVID_meta.txt?dl=0.

## Code availability statement

The GSMR analysis was performed using GCTA 1.93.1beta available at https://cnsgenomics.com/software/gcta/. The genetic correlation analysis was performed using LDSC v1.0.1 available at https://github.com/bulik/ldsc, and HDL v1.3.8 available at https://github.com/zhenin/HDL.

