## Supplementary material for "Genetic and Phenotypic Evidence for the Causal Relationship Between Aging and COVID-19": Covid_supplementary_Oct27.docx

### Supplementary Tables and Figures

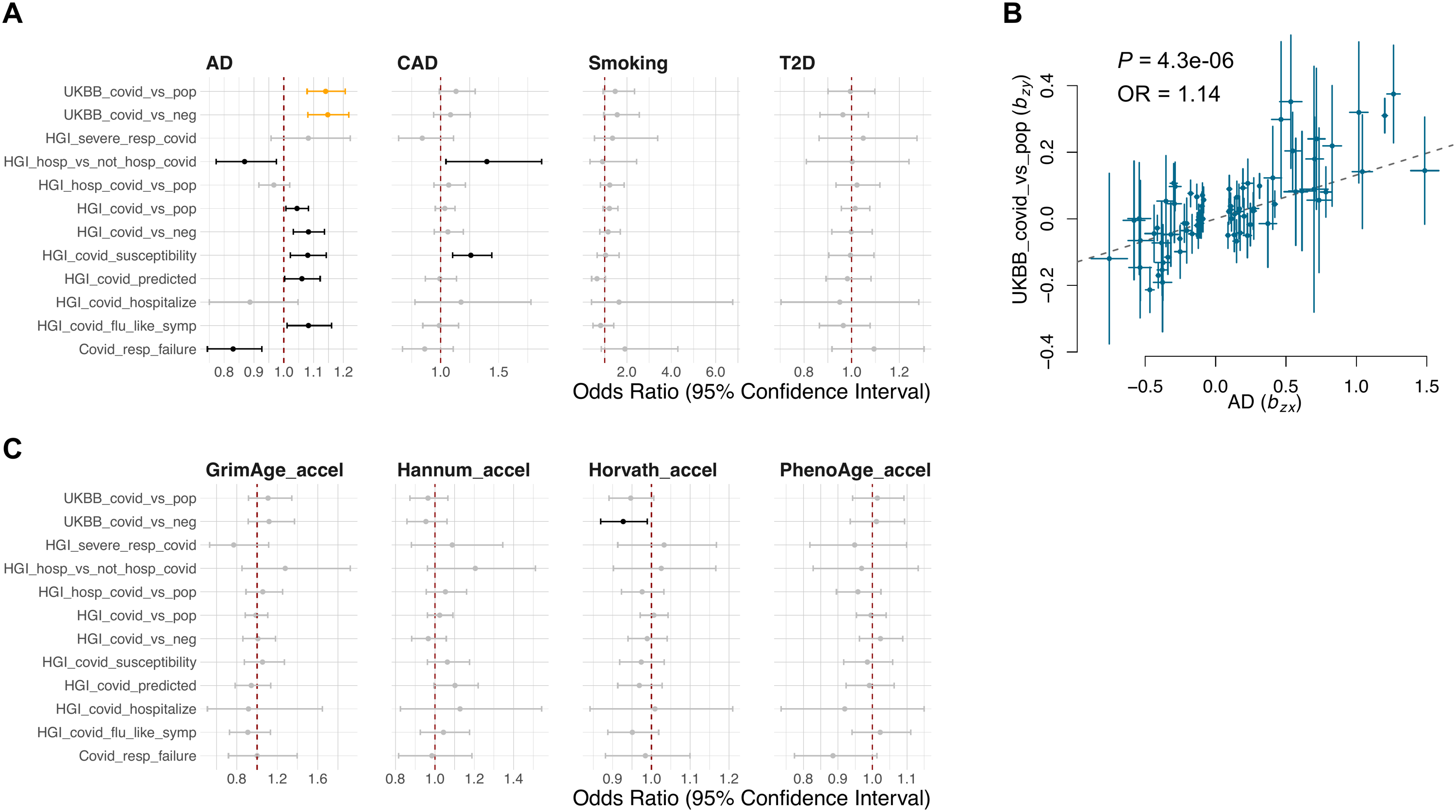
**Figure S1. Mendelian randomization analysis investigating the association of genetically proxied epigenetic age acceleration with the risk of COVID-19.** **A, C.** The figure shows Forest plots and plots of effect sizes showing Mendelian randomization estimates for the causal effect of lifespan-related risk factors **(A)** and epigenetic age acceleration **(C)** on the risk of COVID-19. Error bars show the 95% confidential interval. Significant effects after correcting for 132 tests (P < 0.05/132) are in orange. Nominally significant effects (P < 0.05) are in black. **B.** Plots of effect sizes of all genetic instruments from GWAS for AD (x-axis) versus those for UKBB COVID-19 (y-axis). Error bars represent standard errors.

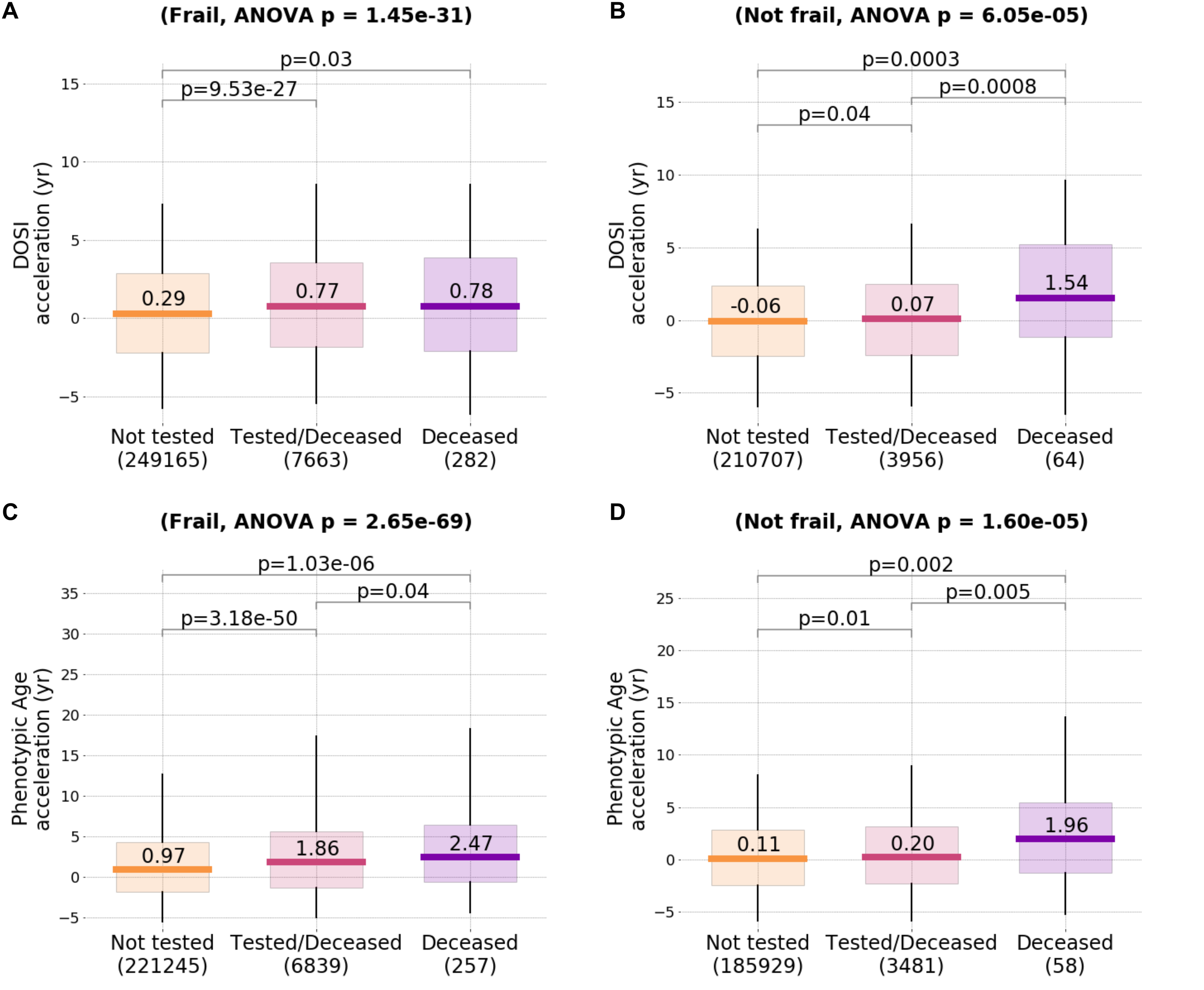

**Figure S2. Biological age acceleration in COVID-19 patients from UKBB cohort.** Box plot showing the distribution of biological age acceleration measured in different groups. Boxes indicate 25%-75% interquartile ranges, and whiskers indicate minimum to maximum. **A, B.** BAA measured by DOSI. **C, D.** BAA measured by Phenotypic Age.

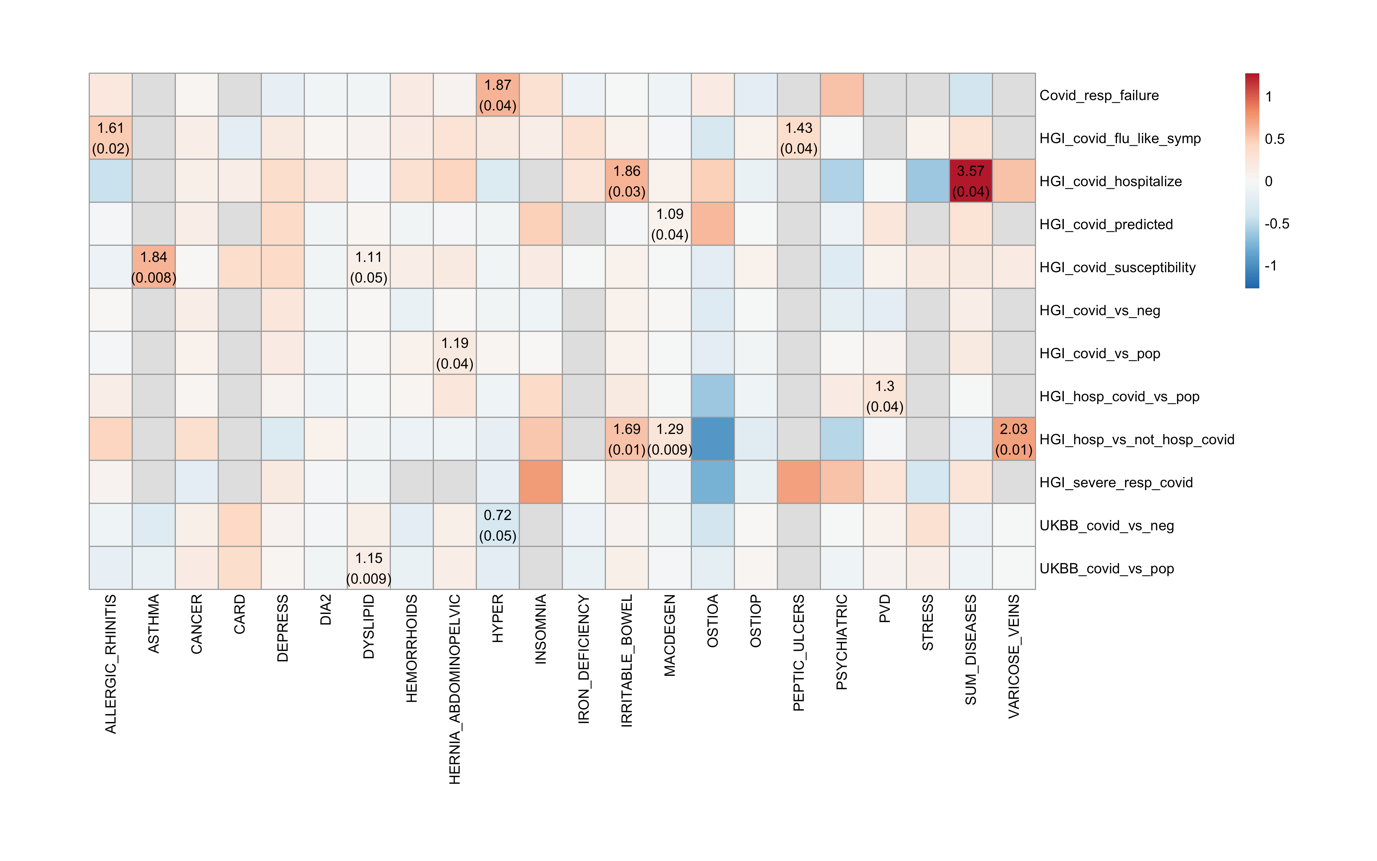

**Figure S3. Putative causal associations between 22 common diseases and the risk of COVID-19.** Shown are the results of GSMR analyses with disease data from a community-based study (GERA). Colors represent the effect sizes (as measured by log odds ratios, log ORs) of diseases on COVID-19, red for risk effects, blue for protective effects, and gray means the effect cannot be estimated due to limited power. Nominally significant effects (*P* < 0.05) are labeled with OR (P-value).

**Figure S4. Twenty bivariate loci identified at genome-wide significance.** **A.** Manhattan plot showing the nominal strength of association −log10(P value) on the y-axis against the chromosomal position of SNPs on the x-axis, where the null hypothesis is no association with healthy aging and COVID-19 infection. **B.** Manhattan plot of the gene-based test as computed by MAGMA test. SNPs were mapped to 18370 protein coding genes. Genome wide significance (red dashed line in the plot) was defined at P = 0.05/18370 = 2.722e-6. **C.** The histogram presenting the summery result per genomic locus. **D.** The histogram displays the proportion of SNPs which have corresponding functional annotation assigned by ANNOVAR. Bars are colored by log2(enrichment) relative to all SNPs in the selected reference panel.

**Table S1. Exposure and outcome trait genetic summary data sources**

| Trait | Abbreviation | Ancestry | N (N_cases_/N_controls_) | Publication (source) | URLs |
| --- | --- | --- | --- | --- | --- |
| Healthspan | Healthspan | European | 300,447 | Zenin *et al.* 2019 *Communications biology* | https://zenodo.org/record/1302861/files/healthspan_summary.csv.gz?download=1 |
| Lifespan | Lifespan | European | 1,012,240 | Timmers *et al.* 2019 *Elife* | https://datashare.is.ed.ac.uk/bitstream/handle/10283/3209/lifegen_phase2_bothpl_alldr_2017_09_18.tsv.gz?sequence=1&isAllowed=y |
| Longevity (age >90th survival percentile) | Longevity | European | 11,262/25,483 | Deelen *et al.* 2019 *Nature Communications* | https://www.longevitygenomics.org/downloads |
| Meta-analysis of Healthspan, Lifespan and Longevity | Healthy aging | European | / | Timmers *et al.* 2020 *Nature Communications* | https://datashare.is.ed.ac.uk/bitstream/handle/10283/3599/timmers2020_healthspan_lifespan_longevity. tsv.gz?sequence=2&isAllowed=y |
| Alzheimer's Disease (late-onset AD) | AD | European | 35,274/59,163 | Kunkle *et al.* 2019 *Nature Genetics* | https://www.niagads.org/igap-rv-summary-stats-kunkle-p-value-data |
| Coronary Artery Disease | CAD | European | 10,801/137,371 | Nelson *et al.* 2017 *Nature genetics* | http://www.cardiogramplusc4d.org/media/cardiogramplusc4d-consortium/data-downloads/ UKBB.GWAS1KG.EXOME.CAD.SOFT.META.PublicRelease.300517.txt.gz |
| Type 2 Diabetes | T2D | European | 62,892/596,424 | Xue *et al.* 2018 *Nature communications* | http://cnsgenomics.com/data.html |
| number of cigarettes smoked per day | Smoking |  | 337,334 | Liu *et al.* 2019 *Nature genetics* | https://conservancy.umn.edu/bitstream/handle/11299/201564/CigarettesPerDay.txt.gz?sequence=31&isAllowed=y |
| Hannum age | Hannum_accel | European | 34,449 | McCartney *et al.* 2020 *BioRxiv* | https://datashare.is.ed.ac.uk/handle/10283/3645 |
| Horvath age | Horvath_accel |  |  |  |  |
| PhenoAge | PhenoAge_accel |  |  |  |  |
| GrimAge | GrimAge_accel |  |  |  |  |
| SARS-COV-2 infection Positive vs. Population | UKBB_covid_vs_pop | European | 1,503 / 457,747 | Genome-Wide Repository of Associations Between SNPs and Phenotypes | https://grasp.nhlbi.nih.gov/downloads/COVID19GWAS/08042020/UKBB_covid19_EUR_080420.txt.gz |
| SARS-COV-2 infection Positive vs. Negative (tested) | UKBB_covid_vs_neg | European | 1,503 / 10,632 | Genome-Wide Repository of Associations Between SNPs and Phenotypes | https://grasp.nhlbi.nih.gov/downloads/COVID19GWAS/08042020/UKBB_covid19_EURtested_080420.txt.gz |
| severe COVID-19 with respiratory failure | Covid_resp_failure | Italy, Spain | 1,610/2,205 | Ellinghaus *et al.* 2020 *New England Journal of Medicine* | www.c19-genetics.eu. |
| ANA2, Hospitalized vs. non-hospitalized | HGI_covid_hospitalize | EUR,  FIN, SAS | 716/616 | The COVID-19 host genetics initiative | https://storage.googleapis.com/covid19-hg-public/20200508/results/COVID19_HGI_ANA2_20200513.txt.gz |
| ANA5, susceptibility (affected vs. population) | HGI_covid_susceptibility | EUR, FIN, SAS, CEU, AFR | 1,678/674,635 | The COVID-19 host genetics initiative | https://storage.googleapis.com/covid19-hg-public/20200508/results/COVID19_HGI_ANA5_20200513.txt.gz |
| ANA7, COVID-19 predicted by flu-like symptoms | HGI_covid_flu_like_symp | EUR | 1294/26,969 | The COVID-19 host genetics initiative | https://storage.googleapis.com/covid19-hg-public/20200508/results/COVID19_HGI_ANA7_20200513.txt.gz |
| very severe respiratory confirmed covid vs. population | HGI_severe_resp_covid | EUR, AMR | 536/329,391 | The COVID-19 host genetics initiative | https://storage.googleapis.com/covid19-hg-public/20200619/results/build_37/COVID19_HGI_ANA_A2_V2_20200701.b37.txt.gz |
| hospitalized covid vs. not hospitalized covid | HGI_hosp_vs_not_hosp_covid | EUR, FIN | 928/2,028 | The COVID-19 host genetics initiative | https://storage.googleapis.com/covid19-hg-public/20200619/results/build_37/COVID19_HGI_ANA_B1_V2_20200701.b37.txt.gz |
| hospitalized covid vs. population | HGI_hosp_vs_pop | EUR, AMR,  SAS, FIN | 3,199/897,488 | The COVID-19 host genetics initiative | https://storage.googleapis.com/covid19-hg-public/20200619/results/build_37/COVID19_HGI_ANA_B2_V2_20200701.b37.txt.gz |
| covid vs. lab/self-reported negative | HGI_covid_vs_neg | EUR, SAS,  AFR, CSA | 3,523/36,634 | The COVID-19 host genetics initiative | https://storage.googleapis.com/covid19-hg-public/20200619/results/build_37/COVID19_HGI_ANA_C1_V2_20200701.b37.txt.gz |
| covid vs. population | HGI_covid_vs_pop | EUR, FIN, ARAB, AMR,  SAS, AFR, CSA | 6,696/1,073,072 | The COVID-19 host genetics initiative | https://storage.googleapis.com/covid19-hg-public/20200619/results/build_37/COVID19_HGI_ANA_C2_V2_20200701.b37.txt.gz |
| predicted covid from self-reported symptoms vs. predicted or self-reported non-covid | HGI_covid_predicted | European | 1,865/29,174 | The COVID-19 host genetics initiative | https://storage.googleapis.com/covid19-hg-public/20200619/results/build_37/COVID19_HGI_ANA_D1_V2_20200701.b37.txt.gz |

**Table S2.** Common diseases in GERA cohort

| **Common Disease** | **Abbreviation** | **N_cases** | **N_controls** |
| --- | --- | --- | --- |
| Asthma | ASTHMA | 10,080 | 51,767 |
| Allergic Rhinitis | ALLERGIC_RHINITIS | 15,166 | 46,681 |
| Cardiovascular Disease | CARD | 16,399 | 45,448 |
| Cancer | CANCER | 18,677 | 43,170 |
| Major Depressive Disorder | DEPRESS | 7,892 | 53,955 |
| Dermatophytosis | / | 8,428 | 53,419 |
| T2D | DIA2 | 7,624 | 54,223 |
| Dyslipidemia | DYSLIPID | 33,024 | 28,823 |
| Hypertensive Disease | HYPER | 31,000 | 30,847 |
| Hemorrhoids | HEMORRHOIDS | 9,898 | 51,949 |
| Hernia Abdominopelvic Cavity | HERNIA_ABDOMINOPELVIC | 6,864 | 54,983 |
| Insomnia | INSOMNIA | 4,346 | 57,501 |
| Iron Deficiency Anemias | IRON_DEFICIENCY | 2,699 | 59,148 |
| Irritable Bowel Syndrome | IRRITABLE_BOWEL | 3,359 | 58,488 |
| Macular Degeneration | MACDEGEN | 4,026 | 57,821 |
| Osteoarthritis | OSTIOA | 22,022 | 39,825 |
| Osteoporosis | OSTIOP | 5,898 | 55,949 |
| Peripheral Vascular Disease | PVD | 4,708 | 57,139 |
| Peptic Ulcer | PEPTIC_ULCERS | 1,004 | 60,843 |
| Psychiatric Disorder | PSYCHIATRIC | 9,394 | 52,453 |
| Acute Reaction to Stress | STRESS | 4,695 | 57,152 |
| Varicose Veins | VARICOSE_VEINS | 2,711 | 59,136 |
